# Analysis of the influence of pollen levels on emergency visits for exacerbations of bronchial asthma in southern Spain

**DOI:** 10.1101/2025.03.24.25324501

**Authors:** Alberto Moreno-Conde, Claudia Rodríguez-Vegas, Jesús Moreno-Conde, Pedro Guardia-Martínez, Angel Vilches-Arenas, Virginia De Luque-Piñana

**Author notes:** **Correspondence to** Alberto Moreno Conde, Unidad de Innovación & Análisis de Datos, Innovation & Data Analysis Unit, Virgen Macarena University Hospital, Av. Dr Fedriani nº9. Seville, 41009, Spain.

## Abstract

**Background:** Epidemiological studies consistently highlight the adverse impact of pollen on asthma exacerbations. This study investigates the correlation between pollen levels and asthma-related Emergency Department visits across four Andalusian provinces, with a combined population of 3.8 million, from 2017 to 2019.

**Materials and methods:** A bidirectional case-crossover design was applied up to 4 lag days to calculate odds ratios (OR) for admissions, adjusting for daily pollen levels and incorporating meteorological factors and personal information as covariates.

**Results:** Significant associations (95% CI) were found between pollen exposure and asthma-related Emergency Department visits, varying by pollen type, region, and lag day. In Almeria, gramineae (1-day lag, OR 1.20); in Huelva, artemisia (1-day lag, OR 1.13), betula (3-day lag, OR 1.26), and olea (1- and 2-day lags, OR 1.20); in Jaén, gramineae (0- and 1-day lags, OR ≈ 9.97), olea (0-4 days, OR ≈ 1.20), and plantago (0-to 2-day lags, OR ≈ 1.20); and in Seville, alternaria (1-4 day lags, OR ≈ 9.96), carex (0-3 day lags, max OR 1.72 at 2 days), olea (0-4 days, OR ≈ 1.20) and urticaceae (same day, OR 1.02).

**Conclusion:** This study provides a comprehensive understanding of pollen’s role in asthma exacerbations, with specific pollen types showing regional and temporal variation. Findings support further research on regional pollen thresholds and geolocation-based strategies to notify asthma patients.

**Summary box:** <u>What do we know about this topic?</u>

<u>How does this study impact our current understanding and/or clinical management of this topic?</u>

## 1. INTRODUCTION

Asthma is a chronic respiratory disease that can affect individuals of all ages. The World Health Organization (WHO) currently estimates 300 million patients with bronchial asthma in the world. Prevalence shows enormous variability between different geographical areas, both in adults (between 0.2% and 21%) and in children (between 2.8% and 37.6%) (1). Differences in asthma prevalence between different studies may be due to a wide variety of causes, including different exposures to risk factors, different diagnostic criteria, ethnic, geographic, socioeconomic, climatic or environmental variations (2).

Several studies have projected substantial increases in aero allergen exposure and associated burden of disease in the USA (3) and Europe (4). Most studies of the effects of pollen have focused on acute effects, as represented in hospital Emergency Department Visits (EDVs) or admissions, occasionally describing inconsistent results or geographical discrepancies (5).

Global Strategy for Asthma Management and Prevention (6) recommends avoidance of outdoors allergens in sensitized patients, such as grass pollen, dust and fungal spores as common exacerbation triggers, specially when pollen and molds counts are highest.

Different systems and tools or applications have been developed to report on the level of different types of pollen, taking into account that each one has a threshold for triggering symptoms. There are networks in several countries such as the USA (7) and Spain (8) that provide results to the public for immediate awareness and utilization. There is a “traffic light notification system” (with colors and alerts) based on previously defined thresholds to inform people of the risk of developing allergy symptoms but does not take into account the geolocation of the patient. It is well known that depending on the area where a patient lives and becomes sensitized, it gives rise to a sensitization profile and a certain pollen threshold is necessary to trigger symptoms (9).

Thus, in a known classic study, about levels of olea europaea pollen and relation with clinical findings, F. Florido et al. observed that level of olive pollen concentration necessary to produce nasal and/or conjunctival symptoms in mono sensitized patients in Jaen was around 400 grains/m3 of Olea Europaea nevertheless classically the threshold described as high for olea Europaea is 200 pollen grains/m3 (10).

By leveraging Geographic Information Systems (GIS), it becomes feasible to gather data concerning the living environment of individuals diagnosed with asthma. The use of tools such as GIS allows the performance of spatial analysis enabling the exploration of various programs, policies, and planning issues in health promotion and public health, including asthma. Several studies pinpoint areas with a high incidence of asthma and compare their environmental conditions with those in regions with lower incidence (11), (12). These tools can be utilized to yield insights into which environmental factors most significantly influence the incidence of asthma in patients, thus allowing patients to avoid risk situations that could cause health problems.

## 2. METHODS

This manuscript describes results from the retrospective analysis of the GeoAsma project. This project aims to advance the study of environmental factors that influence the health of asthmatic patients of the Andalusian Health System (13).

### Location

This was a retrospective study, carried out in the provinces of Seville, Jaen, Huelva and Almeria (other Andalusian provinces were excluded from the conclusions because their pollen stations had less than 300 days of pollen data recordings available per year). In 2017 the study area had 3814022 inhabitants.

### Personal information and health data

The number of EDVs due to asthma exacerbations was obtained from Diraya (Electronic Health Record System of Andalusia). This infrastructure includes data on the geolocation of the residence of the 3.8 million patients it covers in a network of 16 hospitals in the provinces of Almeria, Seville, Huelva and Jaen. For the analysis, we focused on asthma-related emergency visits according to International Classification of Disease V10 codes. The study period ranged from January 1, 2017 to December 31, 2019. Specifically, we utilized information on the patient’s age, sex and date of emergency visit.

### Pollen data

Pollen data was obtained from SEAIC (Spanish Society of Allergology and Clinical Immunology) (14). They have a network of pollen level measurement stations that capture daily presence of some of the best-known types of pollen due to influence on people’s health due to allergies and other health-related problems. The unit of measurement is the number of pollen grains divided by cubic meter (grains/m3). We obtained daily values of 21 different types of pollen particles, detailed in Table 1.

**Table 1:**
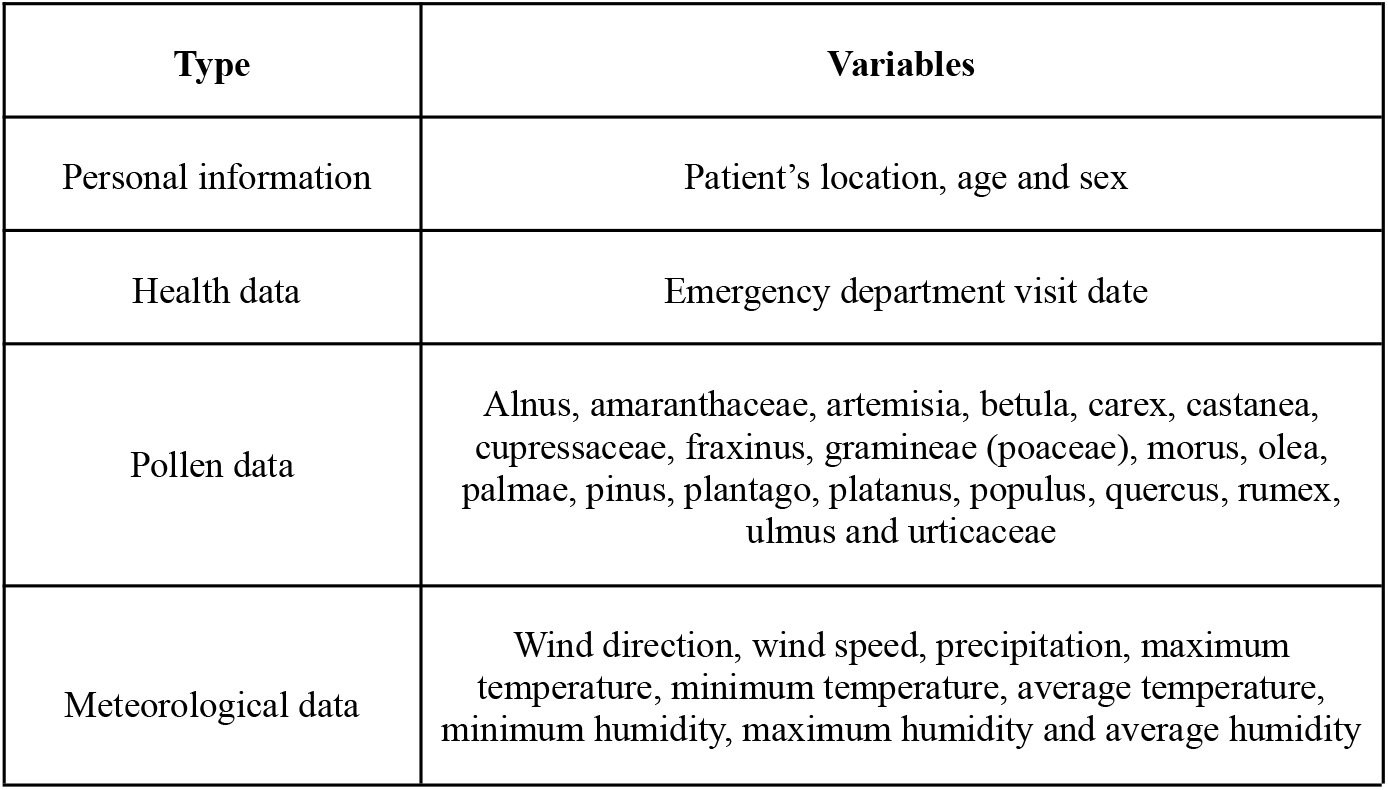
variables taken into account for the study

### Meteorology data

For meteorological variables we incorporated open data from the State Meteorological Agency (AEMET), the Spanish meteorological agency with measurement stations dispersed across the national territory, particularly concentrated in urban centers. The data utilized encompassed daily parameters such as wind (direction and speed), precipitation, temperature (minimum, maximum and average) and humidity (minimum, maximum and average). Table 1 details the list of variables included in this study.

### Analysis techniques

For each province, data was extracted using Ordinary Kriging interpolation technique and a cross-case study was conducted up to lag 4. Using covariates, there were additional controls as follows: altitude, wind direction and wind speed, humidity (minimum, maximum and average), temperature (minimum, maximum and average) and precipitation.

#### Interpolation techniques

In this investigation, we employed an Ordinary Kriging model to project daily pollen and meteorology values across the study area, more information in Annex A1.1. This model utilized point-referenced data from monitoring stations within Andalusia. The Kriging technique integrated observed data and a spatial correlation structure to estimate values at unobserved locations. The data collection process involved gathering point-referenced data from monitoring stations scattered throughout the larger study area, providing insights into observed pollen concentrations at specific locations and times. The model incorporated these data along with prior knowledge, represented as a spatial covariance matrix, to generate predictions for pollen values at unmonitored locations. This method of interpolation was executed drawing upon data from all monitoring stations in Andalusia that were equipped with available data for the respective day. Nevertheless, only four provinces yielded an abundance of over 300 daily records for each year under scrutiny. Consequently, our exploration delves exclusively into the outcomes derived from these particular provinces; Huelva, Seville, Jaen and Almeria.

#### Bidirectional Case-crossover design

A bidirectional case crossover approach was used to study the effects of pollen particles on EDVs due to asthma. Meteorological factors were taken as covariates. For each province each pollen was added to the baseline model to asses de unipollent effect. Only variables that obtained a p-value of at least 0.2 were included in the multipollen model for each province. Using stepwise selection multivariate models were calculated on the same day (lag 0) the consultation took place, the previous day (lag 1) and up to two (lag 2) and three days (lag 3) prior to the visit, recognizing that acute effects of pollen exposure may be immediate or delayed. The case-crossover design was introduced by Maclure in 1991 (15). We used this design to compare pollen particle concentrations during an event with concentrations on a control day. Furthermore, the case-crossover approach allowed the consideration of individual characteristics, such as age or gender, to assess pollen effects among specific subgroups or explore modifications of pollen effects by individual characteristics (16). Control days aligned with the day of the week as the case days, they were selected as 7 days before and after the event, aiming to control time-varying cofactors associated with the day of the week. This design, an adaptation of the case-control study, ensured that each case served as their own control, mitigating the influence of time-invariant subject-specific variables such as gender and age.

## 3. RESULTS

In the studied area the prevalence of asthmatic patients ranges between 8.84% and 11.42% in the studied provinces, furthermore there were 16714 EDVs with asthma as first or secondary diagnosis between 2017 and 2019 (175.30 per 100000 people per year). Mean age was 41.40 years (standard deviation, 23.34 years). 62% of the patients were female. 79% of cases corresponded to adults. Within the studied period, 71% of individuals who experience an EDV did so on a singular occasion. 37% of the visits took place during spring (March 20–June 21). Table 2 details the distribution of population and asthmatic patients in the different provinces.

**Table 2:**
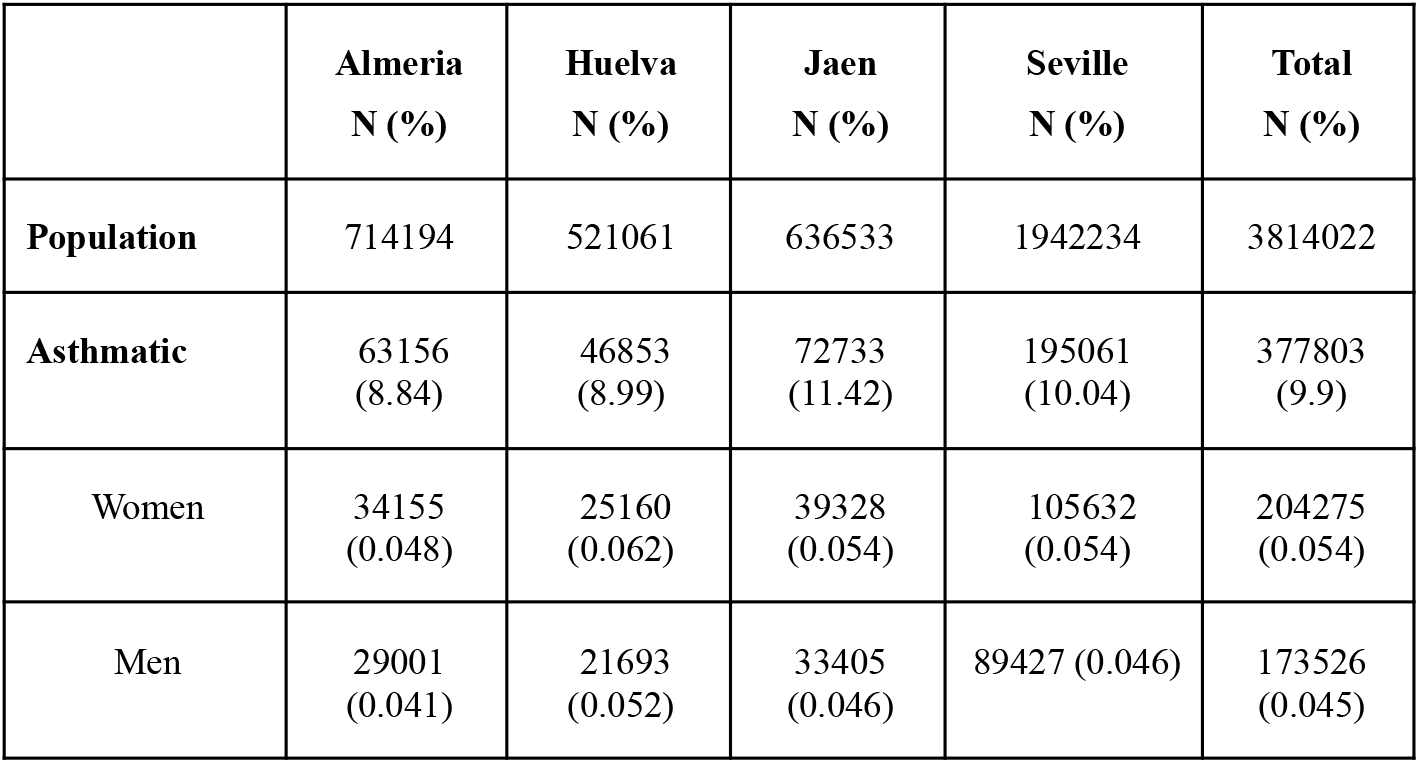
Asthmatic population data for each province from 2017 to 2019

In our population, significant differences among the four provinces studied were observed. Table 3 shows that incidence of EDVs has a distinct temporal distribution pattern, with differences between inland provinces (Jaen and Seville) and coastal provinces (Huelva and Almeria). 37% of the EDVs took place during spring. A peak in attendance during the spring months for inland provinces and during the winter months for coastal ones, was ovserved. In Jaen, we found the peak maximum in the month of May, while in Seville, the peak maximum occurred in April and May.

**Table 3:** EDVs in 2017-2019 for each province

|  | Almería<br>N (%) | Huelva<br>N (%) | Jaen<br>N (%) | Seville<br>N (%) | Total<br>N (%) |
| --- | --- | --- | --- | --- | --- |
| **Season** |  |  |  |  |  |
| Winter<br>(Dec –March) | 1170 (0.31) | 454 (0.34) | 454 (0.19) | 2390 (0.26) | 4468 (0.27) |
| Spring<br>(March –June) | 1084 (0.29) | 397 (0.3) | 1314 (0.55) | 3445 (0.37) | 6240 (0.37) |
| Summer<br>(June –Sept) | 600 (0.16) | 190 (0.14) | 293 (0.12) | 1412 (0.15) | 2495 (0.15) |
| Fall<br>(Sept –Dec) | 862 (0.23) | 302 (0.22) | 350 (0.15) | 1997 (0.22) | 3511 (0.21) |
| **Total** | 3716 (0.06) | 1343 (0.03) | 2411 (0.03) | 9244 (0.05) | 16714 (0.04) |

Using the Variance Inflation Factor (VIF) was analyzed. All VIF values were below 5, indicating that multicollinearity was not a significant issue. Furthermore, these findings were consistent across provinces, confirming that the stability of relationships among variables is maintained across different geographic contexts.

Table 4 shows the OR and their 95%CI for exposure to each pollen with respect to EDVs due to asthma. The pollens that displayed this relationship were:

**Table 4:**
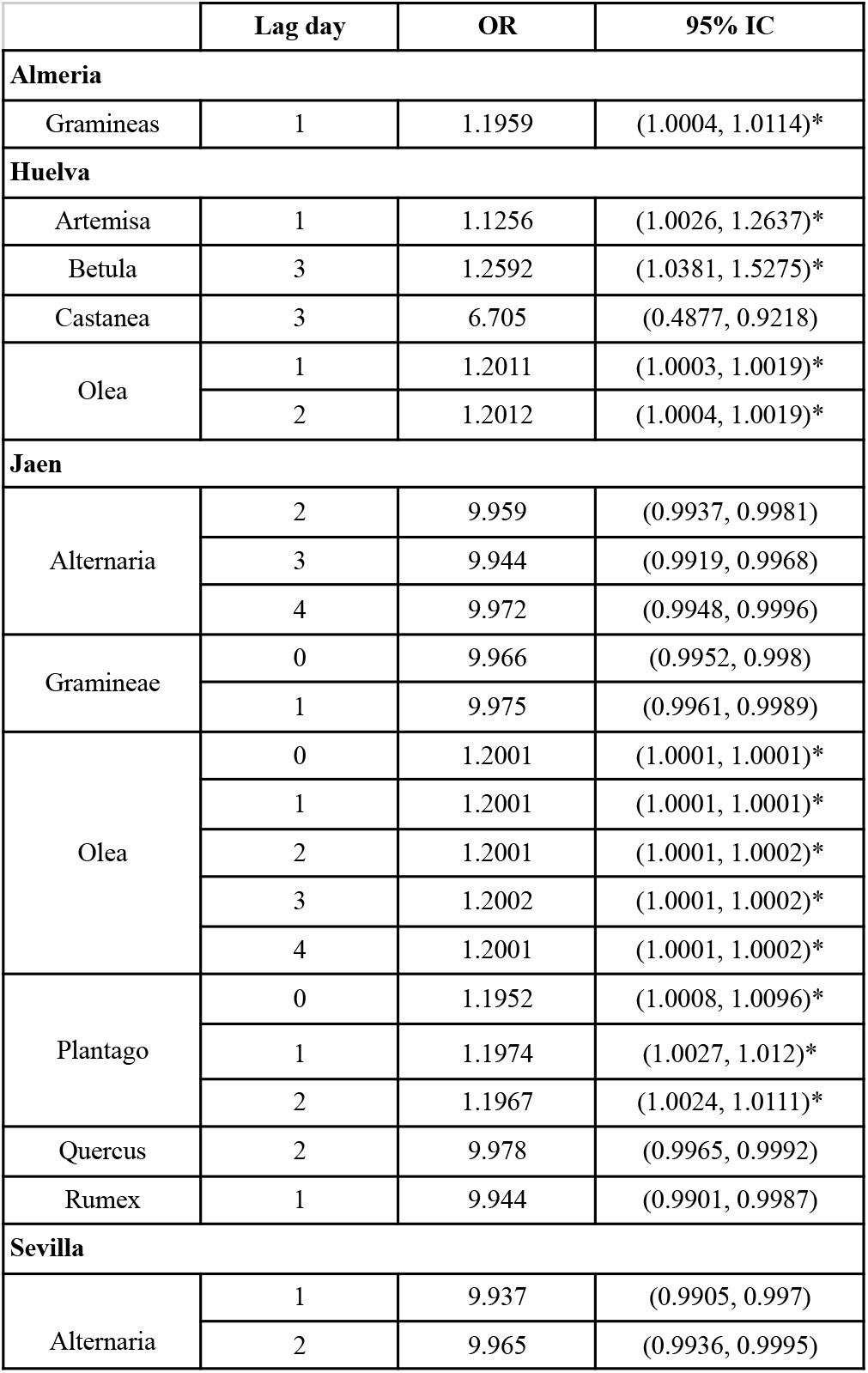
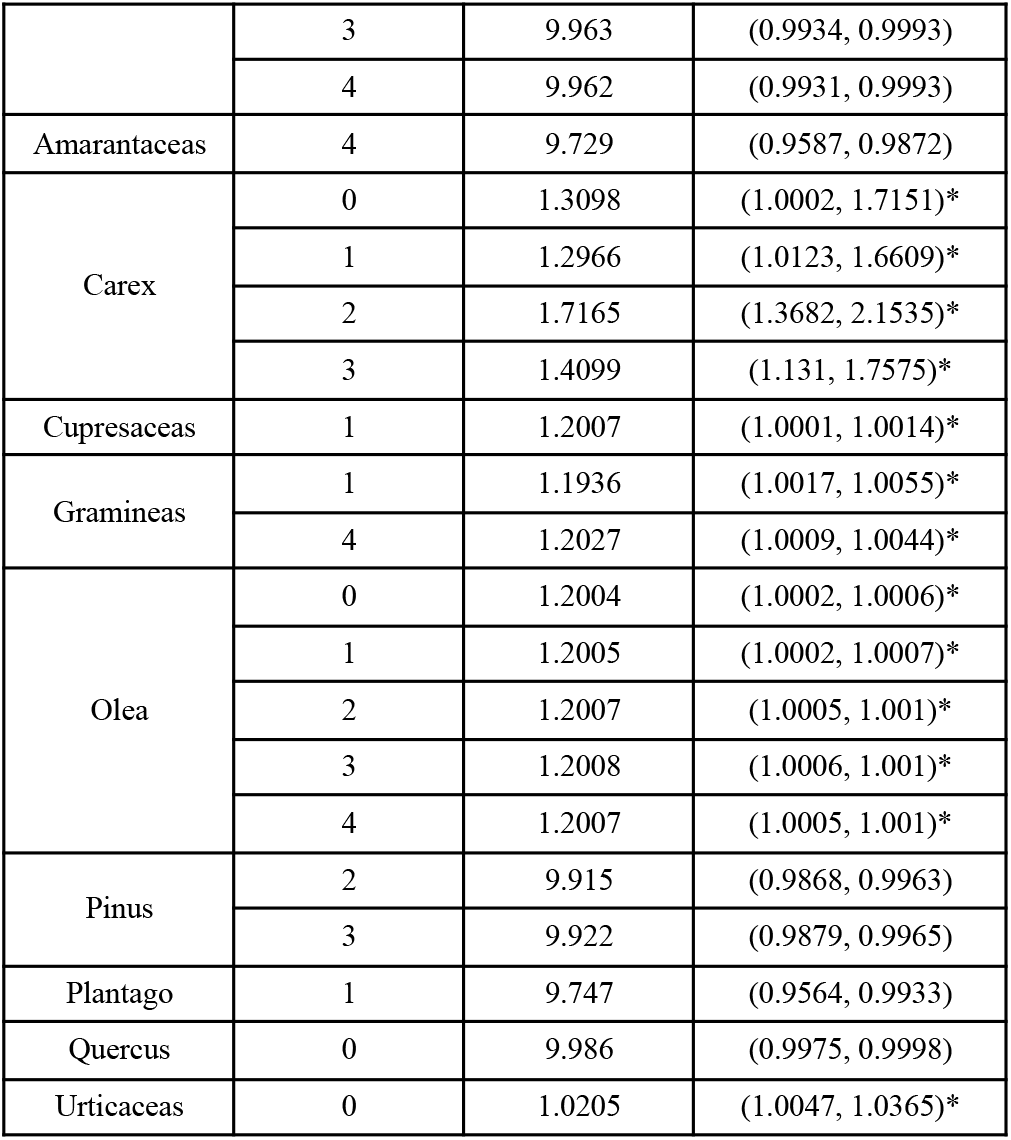
Odds ratios and confidence intervals across lag days that for cases included in the multivariate model according to stepwise selection.

|  | Lag day | OR | 95% IC |
| --- | --- | --- | --- |
| **Almeria** |  |  |  |
| Gramineas | 1 | 1.1959 | (1.0004, 1.0114)* |
| **Huelva** |  |  |  |
| Artemisa | 1 | 1.1256 | (1.0026, 1.2637)* |
| Betula | 3 | 1.2592 | (1.0381, 1.5275)* |
| Castanea | 3 | 6.705 | (0.4877, 0.9218) |
| Olea | 1 | 1.2011 | (1.0003, 1.0019)* |
|  | 2 | 1.2012 | (1.0004, 1.0019)* |
| **Jaen** |  |  |  |
| Alternaria | 2 | 9.959 | (0.9937, 0.9981) |
|  | 3 | 9.944 | (0.9919, 0.9968) |
|  | 4 | 9.972 | (0.9948, 0.9996) |
| Gramineae | 0 | 9.966 | (0.9952, 0.998) |
|  | 1 | 9.975 | (0.9961, 0.9989) |
| Olea | 0 | 1.2001 | (1.0001, 1.0001)* |
|  | 1 | 1.2001 | (1.0001, 1.0001)* |
|  | 2 | 1.2001 | (1.0001, 1.0002)* |
|  | 3 | 1.2002 | (1.0001, 1.0002)* |
|  | 4 | 1.2001 | (1.0001, 1.0002)* |
| Plantago | 0 | 1.1952 | (1.0008, 1.0096)* |
|  | 1 | 1.1974 | (1.0027, 1.012)* |
|  | 2 | 1.1967 | (1.0024, 1.0111)* |
| Quercus | 2 | 9.978 | (0.9965, 0.9992) |
| Rumex | 1 | 9.944 | (0.9901, 0.9987) |
| **Sevilla** |  |  |  |
| Alternaria | 1 | 9.937 | (0.9905, 0.997) |
|  | 2 | 9.965 | (0.9936, 0.9995) |
|  | 3 | 9.963 | (0.9934, 0.9993) |
|  | 4 | 9.962 | (0.9931, 0.9993) |
| Amarantaceas | 4 | 9.729 | (0.9587, 0.9872) |
| Carex | 0 | 1.3098 | (1.0002, 1.7151)* |
|  | 1 | 1.2966 | (1.0123, 1.6609)* |
|  | 2 | 1.7165 | (1.3682, 2.1535)* |
|  | 3 | 1.4099 | (1.131, 1.7575)* |
| Cupresaceas | 1 | 1.2007 | (1.0001, 1.0014)* |
| Gramineas | 1 | 1.1936 | (1.0017, 1.0055)* |
|  | 4 | 1.2027 | (1.0009, 1.0044)* |
| Olea | 0 | 1.2004 | (1.0002, 1.0006)* |
|  | 1 | 1.2005 | (1.0002, 1.0007)* |
|  | 2 | 1.2007 | (1.0005, 1.001)* |
|  | 3 | 1.2008 | (1.0006, 1.001)* |
|  | 4 | 1.2007 | (1.0005, 1.001)* |
| Pinus | 2 | 9.915 | (0.9868, 0.9963) |
|  | 3 | 9.922 | (0.9879, 0.9965) |
| Plantago | 1 | 9.747 | (0.9564, 0.9933) |
| Quercus | 0 | 9.986 | (0.9975, 0.9998) |
| Urticaceas | 0 | 1.0205 | (1.0047, 1.0365)* |

- In Almeria gramineae (lag 0).
- In Huelva artemisa (lag 1), betula (lag 1) and olea (lag 1 - 2) .
- In Jaen olea (lag 0 - 4) and plantago (lag 0 - 2).
- In Sevilla carex and olea (lag 0 - 4), gramineae (lag 0 and lag 4) and urticaceas (lag 0).

Temporal series were calculated, Figure 3 shows the time series corresponding to hospitalizations along with the variables that demonstrate significant importance across multiple lag days. The horizontal lines represent the threshold values for each type of pollen, indicating ‘low’ and ‘high’ levels as defined by the literature for this region’s climate.

**Figure 1.**
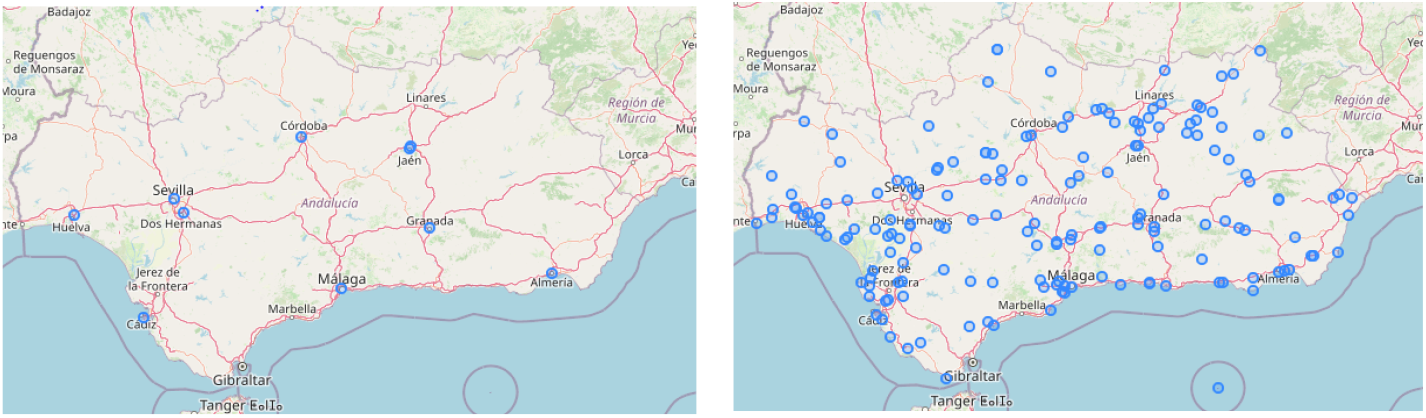
Pollen and meteorological stations providing data for Ordinary Kriging.

**Figure 2.**
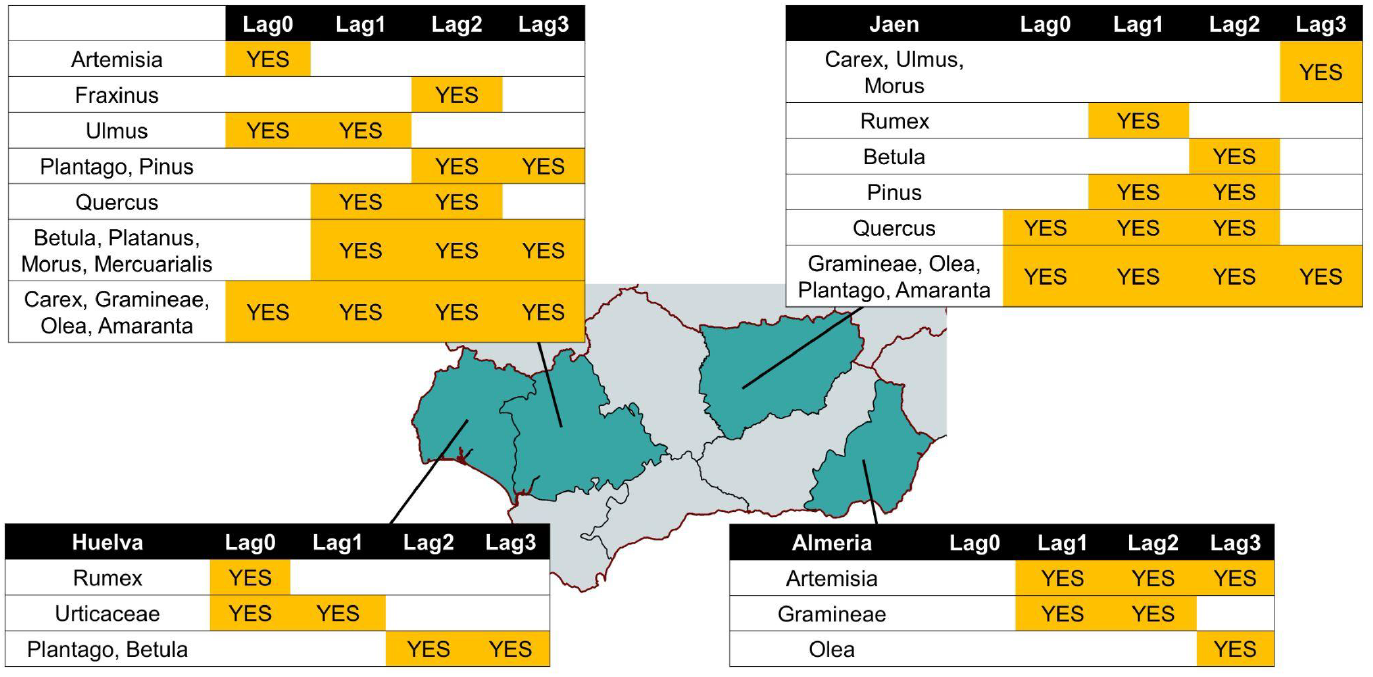
List of pollen particles with p < 0.05 per province depending on the lag days

**Figure 3.**
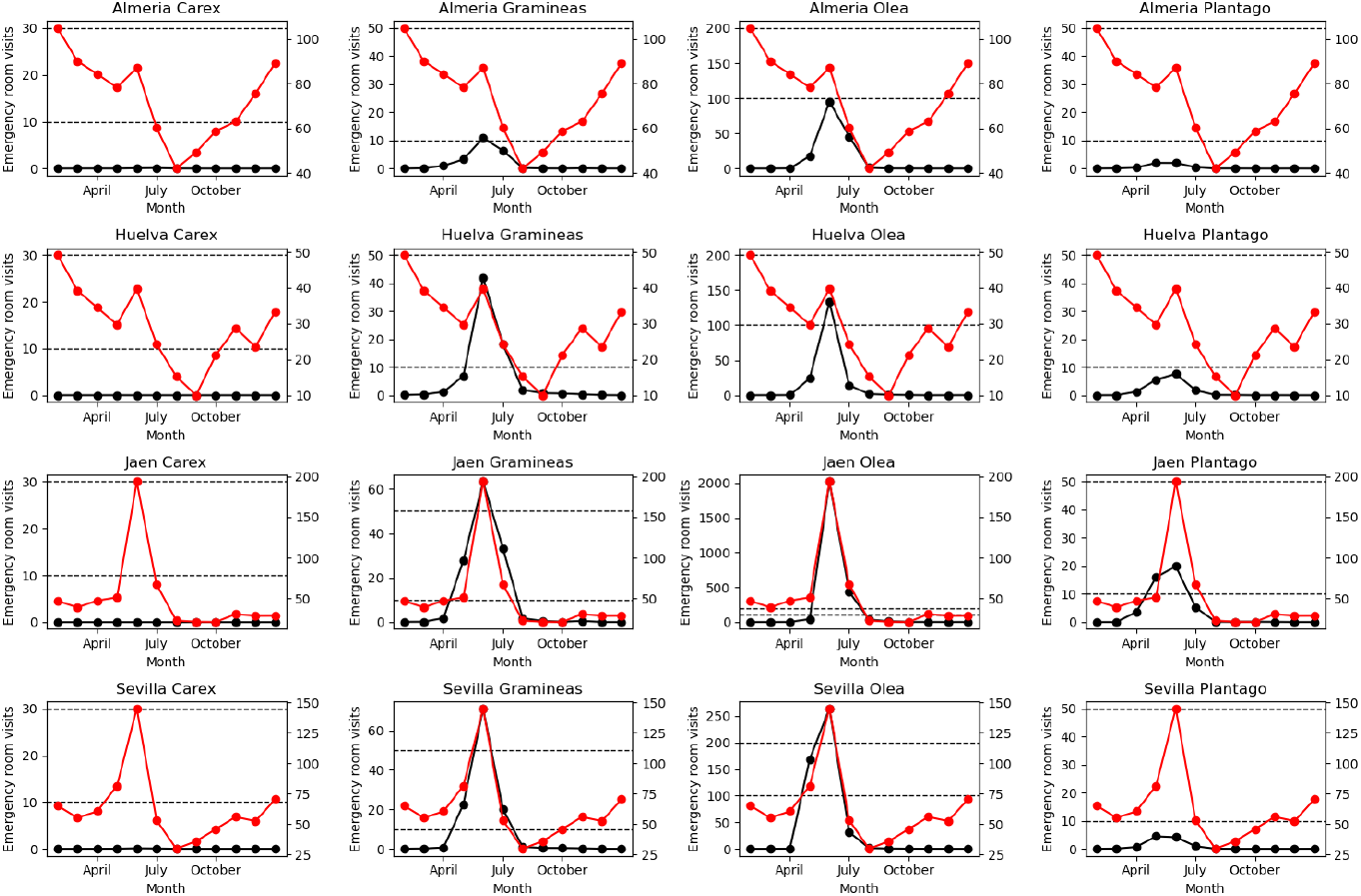
Evolution of the mean monthly values of carex, gramineae, plantago, carex in 2017-2019. Red corresponds with mean monthly EDVS (2017 - 2019) and black with mean monthly pollen value (2017 - 2019)

## 4. DISCUSSION

According to existing literature, prevalence estimates of asthma show significant variability across different geographic areas in both adults (ranging from 0.2% to 21%) and children (ranging from 2.8% to 37.6%). This variability may be attributed to a wide range of causes, including different exposures to risk factors, diverse diagnostic criteria, ethnic, socioeconomic, climatic or environmental variations. Despite this variability, in our population, the prevalence remains within the described range (ranging from 8.8% to 11.4%) matching the prevalence described in other works (2).

In our population, there were 16714 EDVs associated with asthma (175.30 per 100000 people per year). The prevalence described in a study conducted in Spain yielded similar results, with approximately 255.25 EDVs for asthma per 100000 inhabitants per year (18). Other population characteristics such as average age (41.40 years) and gender distribution (54% female patients, 46% male) also align with what is described in the referenced literature.

One aspect worth highlighting, is that only 37% of the visits took place during spring (March 20 - June 21). In coastal provinces, the predominance of EDVs occurred during winter months, possibly corresponding to the coexistence of viral infections or known factors that can precipitate an asthma crisis, such as cold weather. The differing patterns in coastal areas may suggest that in those provinces where pollen plays a less significant role but other allergens such as dust mites or fungi are more prominent, other environmental factors may exert a greater influence. In both Seville and Jaen, the peak of Gramineae and Olea coincides with the times when the highest levels of EDVs occur. Healthcare services must be properly equipped to handle the seasonal surge in demand caused by these patients.

It is striking that this predominance of EDVs in Almeria occurs during winter months, coinciding with a statistically significant relationship between EDVs and the presence of Artemisia pollen. The pollination of this pollen in Almeria during winter months was described years ago, with the highest counts recorded from mid-December to mid-January (19). The observed average for this pollen was 1.57 grains/m3 during winter months, representing the peak compared to other seasons. At this point, two factors likely coincide as predisposing to emergency visits for asthma: low temperatures/winter months, and peak Artemisia pollen count. Further studies would be necessary to corroborate this relationship.

Notably, the pollen level associated with EDVs varies between provinces, a phenomenon previously documented for olive pollen in the study conducted by F. Florido et al. In this study, a threshold of 400 grains per cubic meter was identified as the trigger for rhinitis symptoms. However, the same author acknowledged the necessity for further investigation into this threshold, as it remains unclear whether the level of Olea pollen required to induce asthma symptoms in populations located in different regions is identical. It is imperative to include graphs depicting grass and olive pollen (like figure 3), at least with respect to pollen levels (averaged during spring and maximum value) in Seville and Jaen. These graphs would comprise the two datasets from the temporal series table. The relationship between the increase in the number of admissions and the pollen level threshold described as 400 grains/m3 in Jaen for rhinitis should be explored further. It is crucial to determine whether this threshold applies equally to asthma symptoms, as this information is vital for various applications.

Different thresholds are used in daily pollen concentrations to study the allergenic potential of different species of pollen grains. Even within the established ranges in the same country, these ranges do not always coincide. For example, the REA considers a moderate range of birch pollen to be from 30 to 50 grains/m3 (20), while according to the SEAIC it is from 40 to 80 grains/m3 (21). When considering other European countries, different results are obtained as well. For instance, according to the Federal Office of Meteorology and Climatology MeteoSwiss, a moderate range of birch pollen is from 11 to 69 grains/m3 (22).

Understanding the disparities between provinces is essential for comprehensive analysis and effective decision-making. For example, we found statistical significance of plantago in Jaen. However, as shown in Figure 3, the monthly mean value of this pollen was considered low according to the threshold tables defined by both the REA and SEAIC.

It is worth noting the increased risk of EDVs observed in the province of Seville for birch pollen. Traditionally, this pollen has not been considered relevant in our area (although it is in Northern Europe), as seen in previous studies, with a very low frequency of sensitization in patients with rhinitis or asthma compared to other types of pollen. However, it is true that an increase has been observed in patients with rhinitis sensitized to birch pollen, from 0.5 in 2005 to 1.6 in 2015 in Spain (23). Further studies are needed to define the real significance of this increased risk, whether it is due to birch pollen itself, or if the classic priming effect after weeks of exposure to high levels of other types of pollen could be influencing it.

The results obtained provide a comprehensive understanding of pollen’s influence in four provinces within the Andalusian region, aiming to formulate customized strategies and recommendations based on the associated risks. This information holds potential relevance in offering valuable insights into the patient environment for clinicians and patients in the analyzed areas. Designing strategies that consider the geolocation of patients is essential, as pollen level thresholds cannot be universally applied across different provinces. Monitoring systems and tools to report on the level of different types of pollen need to take into account these differences to provide appropriate support to patients and general population. The results of these analyses will be implemented in a decision support system for professionals and a mobile app for patients that will include real time notifications according to the environmental conditions. The impact of adopting these systems in clinical practice will be evaluated through a pilot with 214 patients with a follow-up period of 12 months with asthma in 8 Andalusian hospitals (13). We advocate for further research on the defined thresholds for each type of pollen, preferably conducted on a smaller regional scale than previously proposed. Such research could prove instrumental in controlling asthma exacerbations.

Previous studies have identified a limitation, which we also face, concerning the variability in defining an exacerbation and diagnosing bronchial asthma. This factor could potentially impact the results obtained, although our findings align with asthma prevalence values, population characteristics, and emergency department attendance rates reported in other studies. Another constraint linked to the interpolation method of pollen values across provinces is the paucity of monitoring stations throughout the study area. Furthermore, a limitation arising from the study design is the absence of information regarding the use of rescue medication, which could substantially influence a patient’s decision to seek emergency care.

## Ethical approval

This research has been approved by the Andalusian Biomedical Research Ethics Committee Coordinator under protocol number 2403-N-20. The committee granted exemption from informed consent for retrospective studies based on the safeguard mechanisms defined in Spanish Law 3/2018 on personal data protection and digital rights, which mandates technical and functional separation between researchers and technicians responsible for patient data pseudonymization.

## Author’s contribution

AMC and VDP defined the analysis and leaded the manuscript writing. CRV performed the analysis described in the methodology section and drafted the word. AVA and PGM provided guidance about how to perform the analysis and discussion. JMC implemented the database and underlying technology. All authors have reviewed and approved the final version of the article.

## Data Availability

All data produced in the present study are available upon approval from the Andalusian Health Service Access Committee.

## Abbreviations and acronyms

EDV: Emergency Department Visit
WHO: The World Health Organization
the USA: the United States of America
GIS: Geographic Information Systems
SEAIC: Spanish Society of Allergology and Clinical Immunology
AEMET: State Meteorological Agency AEMET
REA: Spanish Aerobiology Network.
(VIF): Variance Inflation Factor

